# Are Suboptimal *in utero* Conditions Associated with Obesity and Cardiometabolic Risk Factors in Caucasian children?

**DOI:** 10.1101/2020.06.18.20132837

**Authors:** Soraya Saidj, Melanie Henderson, Stephanie-May Ruchat, Gilles Paradis, Andraea Van Hulst, Michael Zappitelli, Marie-Eve Mathieu

**Author notes:** **Corresponding author:** Marie-Eve Mathieu, École de kinésiologie et des sciences de l’activité physique, Université de Montréal, CEPSUM 2100, boul. Édouard-Montpetit, Bureau 8202, Montréal (Canada) H3T 1J4. **Authors disclosure statement**No competing financial interests exist.

## Abstract

**Objective:** To explore the association between *in utero* exposure to suboptimal gestational factors (SGFS; gestational diabetes mellitus, hypertensive disorders during pregnancy, maternal smoking during pregnancy), childhood obesity, and cardiometabolic risk factors.

**Methods:** Data were obtained from the “QUebec Adiposity and Lifestyle InvesTigation in Youth” longitudinal study (QUALITY) composed of 630 Caucasian children recruited at age 8–10 (first visit V1; n=619) and followed-up 2 years later (second visit V2; n=557). Multivariate logistic regression models were used.

**Results:** In the cohort, maternal smoking during pregnancy was associated with higher odds of obesity (OR1=2.00, 95% CI 1.25-3.20, OR2=2.29, 95% CI 1.26-4.16) at both visit and with higher odds of high waist circumference (OR1=1.96, 95% CI 1.24-3.1) at V1. Hypertensive disorders during pregnancy were associated with higher odds of obesity (OR1=2.37, 95% CI 1.17-4.80) at visit 1. Joint exposure to SGFS was associated with higher odds of: high waist circumference (OR1=1.42, 95% CI 1.06-1.91) at V1, obesity (OR2= 1.63, 95% CI 1.153-2.32) at V2 and low levels of HDL cholesterol (OR2=1.45, 95% CI 1.04-2.03) at V2. Analysis stratification by sex revealed that boys were more adversely affected by exposure to combined SGFS while girls were more affected by exposure to maternal smoking during pregnancy.

**Conclusion:** *In utero* exposure to an independent or combined SGFS is associated with adverse adipose and cardiometabolic profiles in children, with sex specificities.

## Introduction

Childhood obesity has been attributed to an interaction of genetic and non-genetic factors such as a shift toward a less active lifestyle^1^ and low-nutrient, energy-dense diets^2^. Prenatal and possibly pre-conceptual factors may also play a role. According to Barker’s theory of “*Developmental Origins of Health and Disease*” (DOHaD)^3^, an adverse environment early during development, particularly during intrauterine life, can result in permanent changes in fetus physiology and metabolism, with higher risks of later cardiometabolic diseases. Offspring exposed to gestational diabetes mellitus (GDM) are more likely to be overweight or obese^4^ and to present components of the metabolic syndrome during childhood^5^ and early adolescence^6^. According to Clausen *et al*. (2009)^7^, the risk of metabolic syndrome in adulthood is four folds higher in a population exposed to GDM when compared with the general population. Similarly, offspring exposed to maternal hypertension during pregnancy are more likely to have high blood pressure during childhood^8^ and adolescence^9^ and to be overweight or obese in childhood^10^. A third factor of interest is maternal smoking during pregnancy. Offspring exposed to maternal smoking during pregnancy are more susceptible to be obese during childhood^11^, adolescence and young adulthood^12^, and to display higher levels of triglycerides and a lower level of high-density lipoprotein (HDL)^13^ compared to the non-exposed population.

The association between suboptimal gestational factors (SGFS) (GDM, hypertensive disorders of pregnancy, maternal smoking during pregnancy), obesity and cardiometabolic risk factors in children between age 8–10 and 10–12 years is to be confirmed. In addition, the association between joint exposure to SGFS and obesity, as well as cardiometabolic risk factors during childhood remains to be established. This study aimed to explore, in a cohort of Caucasian children aged 8–10 (late childhood) at baseline and followed at 10–12 years of age (pubertal onset), the association between independent and joint exposure to SGFS, obesity and cardiometabolic risk factors.

## Methods

### Population

This prospective study used data from the QUebec Adiposity and Lifestyle InvesTigation in Youth (QUALITY) study designed to increase understanding of the natural history of excess body weight and its associated cardiometabolic consequences in Caucasian youth at risk for the development of obesity (http://www.etudequalitystudy.ca)^14^. Data from a sample of 630 children aged 8–10 years at baseline [Visit 1 (V1); 2005–2008] of which 564 were followed 2 years later [Visit 2, 2007-2011 (V2)] were used. Inclusion criteria were for children to have at least one obese biological parent (i.e., body mass index [BMI] ≥ 30 kg/m^2^ and/or a waist circumference of at least 88 cm for women or 102 cm for men). Exclusion criteria for children were to have type 1 or 2 diabetes; to have antihypertensive medications or steroids use; to follow a very strict diet (i.e. <600 kcal/day); to have a previous diagnosis of a serious illness, a psychological condition, or a cognitive disorder that precluded participation. The current analysis is restricted to children who were reported to be non-exposed to type 1 or 2 diabetes during pregnancy (pre-gestational diabetes) considering the differences in medical care, maternal and fetal outcomes with these two medical conditions^15^. The QUALITY study was approved by the Ethics Review Boards of the Sainte-Justine University Hospital Center and Québec Heart and Lung Institute. Written informed parental consent and child assent were obtained.

### Suboptimal gestational factors

Data on maternal health conditions during pregnancy (i.e. GDM, hypertensive disorders during pregnancy, and maternal smoking during pregnancy) were collected using a self-administered questionnaire to parents, an approach known to be valid^16^. In the questionnaire, parents had to answer “yes” or “no” or “I don’t know” to the following questions, “Was the mother diagnosed for gestational diabetes during her pregnancy?”, “Was the mother diagnosed for hypertension during pregnancy?” and “Did the mother smoke during her pregnancy?”. Joint exposure to SGFS was defined as the number of SGFS to which the child was exposed to during pregnancy (0, 1, 2 or 3).

### Anthropometric measures and obesity diagnosis

All anthropometric measures were obtained using standardized methods. Height was measured twice using a calibrated stadiometer (Ibiom, model 600, Ibiom Instruments Ltée, Sherbrooke, Quebec, Canada at Sainte-Justine Hospital and Medical Scales and Measuring Devices Seca Corp., Hanover, Maryland, USA at Quebec Heart and Lung Institute). Body weight was measured with a calibrated electronic balance at Sainte-Justine’s hospital (Cardinal Detecto, 758C Series, Cardinal Scale Manufacturing Co., Webb City, Missouri, USA) and at the Laval hospital (Tanita, model TBF-300A, Arlington Heights, Illinois, USA). Waist circumference was measured twice to the closet 0.1cm at the midpoint between the lower margin of the least palpable rib and the top of the iliac crest using a calibrated length measuring tape. High waist circumference was determined at or above the 90^th^ percentiles using the International Diabetes Federation (IDF) consensus statement^17^.

Obesity definition is based on BMI for age and sex. BMI percentile for age and sex is computed and interpreted using the Centers for Disease Control and Prevention age and sex-specific reference norms^18^. Obesity is defined as a BMI at or above the 95th percentile for children and teens of the same age and sex^19^.

### Cardiometabolic risk factors

The current definition of metabolic syndrome in childhood by the International Diabetes Federation consensus statement of 2007 was used^17^. A definition still used as a reference today^20^. It proposes that metabolic syndrome in children greater than 10 years of age can be diagnosed by the presence of central abdominal obesity and two or more clinical features: elevated triglycerides, low level of HDL, hypertension, and increased plasma glucose^17^. Blood pressure was measured on the right arm, using a size-appropriate cuff, after a 5-minute rest, in a seated position using an oscillometric instrument (Dinamap XL, model CR9340, Critikon Company, FL, USA). Five consecutive readings were recorded and the mean value of the last three readings was used to define systolic and diastolic blood pressure. Hypertension was diagnosed if systolic ≥130 and diastolic ≥85 mm Hg^17^. Blood samples were obtained by venipuncture after an overnight fast by a trained nurse. Analyses were performed in batches at Sainte-Justine hospital’s clinical biochemistry laboratory twice monthly. Fasting hyperglycemia was diagnosed if fasting glycemia>=5.6 mmol/L^17^. Adverse lipid profile was diagnosed if HDL was <1.03 mmol/L or if triglycerides>=1.7 mmol/L^17^.

### Statistical analysis

The sample was described using means and percentages. Multivariate logistic regression was used to investigate the association between each dichotomous independent variable (GDM, hypertensive disorders during pregnancy and maternal smoking during pregnancy), obesity (dependent variable) as well as cardiometabolic risk factors (Waist circumference, levels of HDL cholesterol, fasting glycemia, triglycerides, and blood pressure; dependent variables). Multivariate logistic regression was used to investigate the association between the independent categorical variable “joint exposure to SGFS” (0 factor, 1 factor, 2 factors and 3 factors), obesity as well as cardiometabolic risk factors (dependent variables). Analyses were adjusted for parental education, and models for systolic as well as diastolic blood pressure were adjusted for height at outcome measurement. Data were analyzed using SPSS version 24 (IBM Corp. 2012. IBM SPSS Statistics for Windows, Armonk, NY, USA), and covariate-adjusted associations are reported.

### Adjusted variables

Parental education^21^ could be associated with obesity in children and was controlled for in the analysis. Data about parental education were collected using a self-administered questionnaire to parents, an approach known to be valid^16^.

## Results

Age, sex, anthropometric measures, exposure to SGFS and cardiometabolic risk factors for V1 and V2 are described in Table 1. From the initial sample of 630 (V1) and 564 (V2), 11 (V1) and 7 (V2) children were excluded from the study due to reported diabetes in the mother before pregnancy. The analytic samples of 619 for V1 and 557 for V2 were used. Age, gender, parent’s education level, did not differ between children who were present at both visits and children lost to follow up (data not presented).

**Tables 1:**
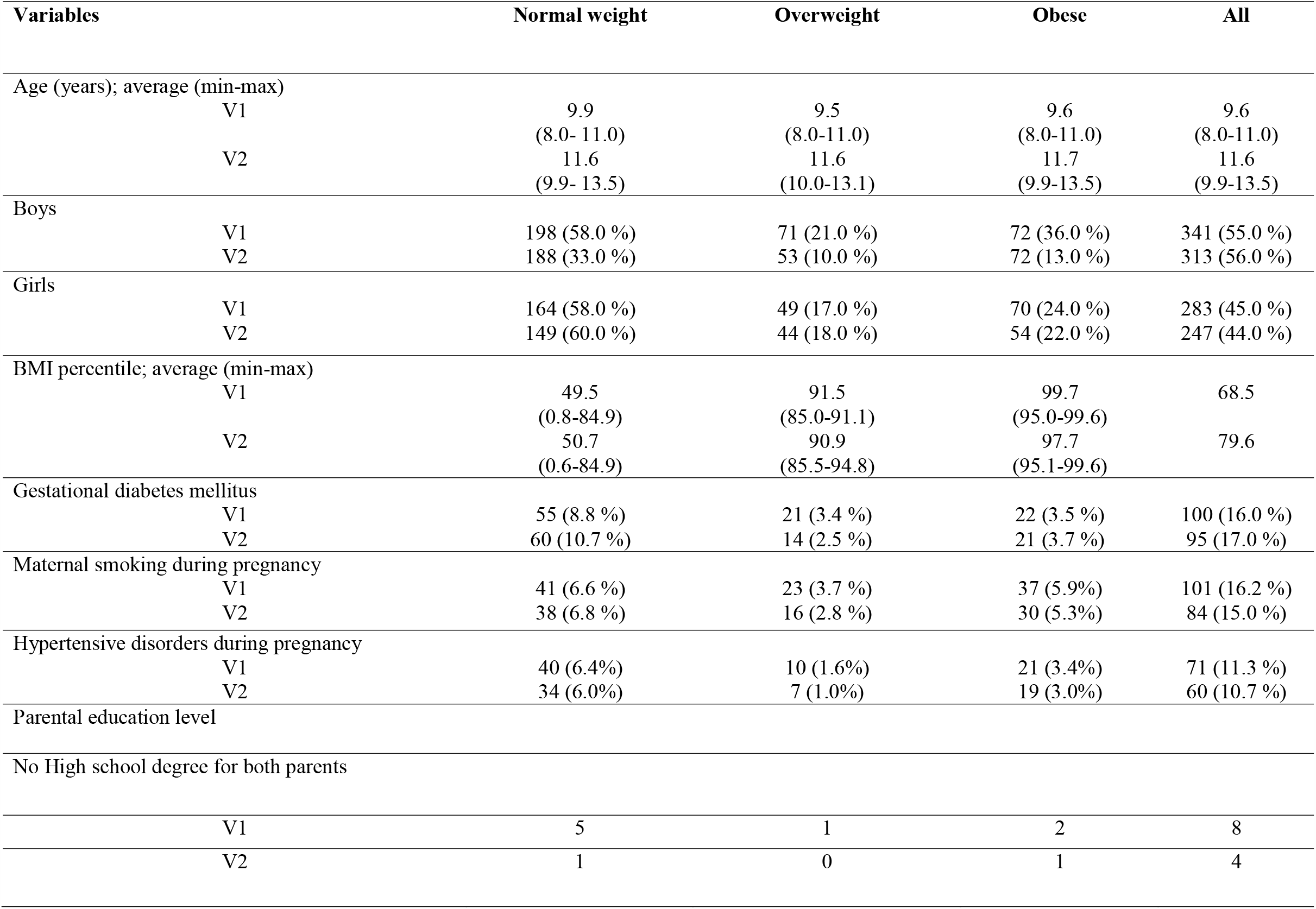
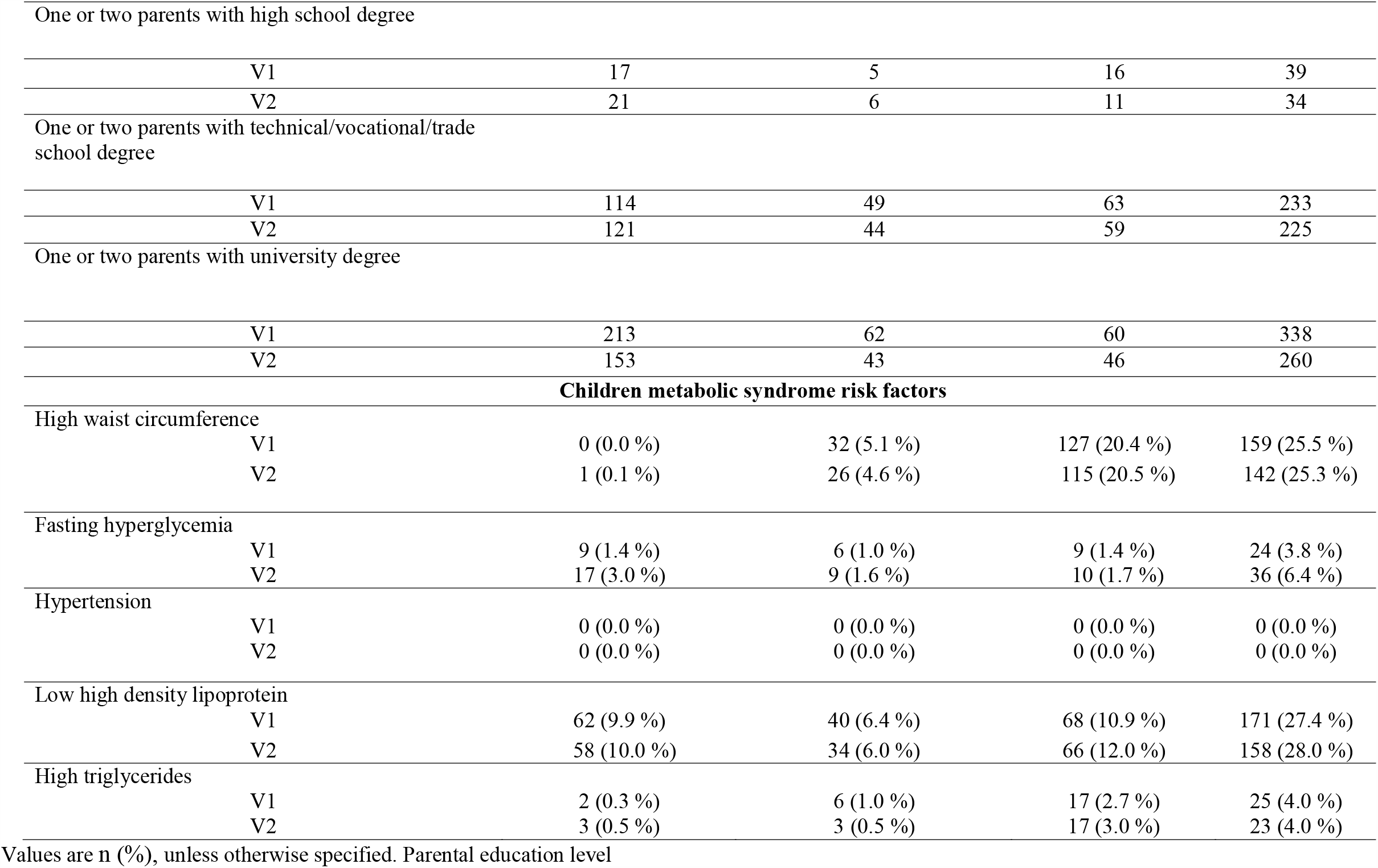
Study sample characteristics at 8-10 years (V1) and 10-12 years (V2)

| Variables | Normal weight | Overweight | Obese | All |
| --- | --- | --- | --- | --- |
| Age (years); average (min-max) |  |  |  |  |
| V1 | 9.9<br>(8.0- 11.0) | 9.5<br>(8.0-11.0) | 9.6<br>(8.0-11.0) | 9.6<br>(8.0-11.0) |
| V2 | 11.6<br>(9.9- 13.5) | 11.6<br>(10.0-13.1) | 11.7<br>(9.9-13.5) | 11.6<br>(9.9-13.5) |
| Boys |  |  |  |  |
| V1 | 198 (58.0 %) | 71 (21.0 %) | 72 (36.0 %) | 341 (55.0 %) |
| V2 | 188 (33.0 %) | 53 (10.0 %) | 72 (13.0 %) | 313 (56.0 %) |
| Girls |  |  |  |  |
| V1 | 164 (58.0 %) | 49 (17.0 %) | 70 (24.0 %) | 283 (45.0 %) |
| V2 | 149 (60.0 %) | 44 (18.0 %) | 54 (22.0 %) | 247 (44.0 %) |
| BMI percentile; average (min-max) |  |  |  |  |
| V1 | 49.5<br>(0.8-84.9) | 91.5<br>(85.0-91.1) | 99.7<br>(95.0-99.6) | 68.5 |
| V2 | 50.7<br>(0.6-84.9) | 90.9<br>(85.5-94.8) | 97.7<br>(95.1-99.6) | 79.6 |
| Gestational diabetes mellitus |  |  |  |  |
| V1 | 55 (8.8 %) | 21 (3.4 %) | 22 (3.5 %) | 100 (16.0 %) |
| V2 | 60 (10.7 %) | 14 (2.5 %) | 21 (3.7 %) | 95 (17.0 %) |
| Maternal smoking during pregnancy |  |  |  |  |
| V1 | 41 (6.6 %) | 23 (3.7 %) | 37 (5.9%) | 101 (16.2 %) |
| V2 | 38 (6.8 %) | 16 (2.8 %) | 30 (5.3%) | 84 (15.0 %) |
| Hypertensive disorders during pregnancy |  |  |  |  |
| V1 | 40 (6.4%) | 10 (1.6%) | 21 (3.4%) | 71 (11.3 %) |
| V2 | 34 (6.0%) | 7 (1.0%) | 19 (3.0%) | 60 (10.7 %) |
| Parental education level |  |  |  |  |
| No High school degree for both parents |  |  |  |  |
| V1 | 5 | 1 | 2 | 8 |
| V2 | 1 | 0 | 1 | 4 |
| One or two parents with high school degree |  |  |  |  |
| V1 | 17 | 5 | 16 | 39 |
| V2 | 21 | 6 | 11 | 34 |
| One or two parents with technical/vocational/trade school degree |  |  |  |  |
| V1 | 114 | 49 | 63 | 233 |
| V2 | 121 | 44 | 59 | 225 |
| One or two parents with university degree |  |  |  |  |
| V1 | 213 | 62 | 60 | 338 |
| V2 | 153 | 43 | 46 | 260 |
| **Children metabolic syndrome risk factors** |  |  |  |  |
| High waist circumference |  |  |  |  |
| V1 | 0 (0.0 %) | 32 (5.1 %) | 127 (20.4 %) | 159 (25.5 %) |
| V2 | 1 (0.1 %) | 26 (4.6 %) | 115 (20.5 %) | 142 (25.3 %) |
| Fasting hyperglycemia |  |  |  |  |
| V1 | 9 (1.4 %) | 6 (1.0 %) | 9 (1.4 %) | 24 (3.8 %) |
| V2 | 17 (3.0 %) | 9 (1.6 %) | 10 (1.7 %) | 36 (6.4 %) |
| Hypertension |  |  |  |  |
| V1 | 0 (0.0 %) | 0 (0.0 %) | 0 (0.0 %) | 0 (0.0 %) |
| V2 | 0 (0.0 %) | 0 (0.0 %) | 0 (0.0 %) | 0 (0.0 %) |
| Low high density lipoprotein |  |  |  |  |
| V1 | 62 (9.9 %) | 40 (6.4 %) | 68 (10.9 %) | 171 (27.4 %) |
| V2 | 58 (10.0 %) | 34 (6.0 %) | 66 (12.0 %) | 158 (28.0 %) |
| High triglycerides |  |  |  |  |
| V1 | 2 (0.3 %) | 6 (1.0 %) | 17 (2.7 %) | 25 (4.0 %) |
| V2 | 3 (0.5 %) | 3 (0.5 %) | 17 (3.0 %) | 23 (4.0 %) |

Values are n (%), unless otherwise specified. Parental education level

### Visit 1 and Visit 2

In the cohort (boys and girls combined) at age 8–10 (V1), maternal smoking during pregnancy was positively associated with 100% higher odds of obesity (OR=2.00, 95% CI 1.25-3.20) (Table 2) and 96% higher odds of high waist circumference (OR=1.96, 95% CI 1.29-5.17) (Table 3). Joint exposure to SGFS was associated with 37% higher odds of high waist circumference (OR=1.37, 95% CI 1.01-1.86) at V1 (Table 3). At age 10–12 years, maternal smoking during pregnancy was positively associated with 129% higher odds of obesity (OR=2.29, 95% CI 1.26-4.16) (Table 4) (Figure1). Joint exposure to SGFS was positively associated with 63% higher odds of obesity (OR=1.63, 95% CI 1.15-2.32) (Table 4) and 45 % higher odds of low levels of HDL (OR=1.45, 95% CI 1.04-2.03) (Table 5). Comparison between odd ratios of significant associations which persisted between the age of 8–10 years (V1) and age of 10–12 years (V2), showed no significant difference.

**Table 2:** Adjusted multivariate logistic regression model assessing the association between anthropometric measures and suboptimal gestational factors in QUALITY cohort and boys and girls at age 8-10 years (Visit1)

| Suboptimal gestational condition | Population | Overweight |  | Obesity |  |
| --- | --- | --- | --- | --- | --- |
|  |  | OR | 95% CI | OR | 95% CI |
| Gestational diabetes mellitus | All | 1.14 | 0.65-1.92 | 0.90 | 0.53-1.52 |
|  | Girls | 1.21 | 0.75-1.93 | 0.55 | 0.49-2.62 |
|  | Boys | 0.91 | 0.61-1.38 | 0.82 | 0.41-1.61 |
| Hypertensive disorders during pregnancy | All | 0.98 | 0.73-1.33 | 1.50 | 0.86-2.62 |
|  | *Girls* | 0.48 | 0.14-1.67 | 0.77 | 0.29-2.00 |
|  | *Boys* | 0.81 | 0.34-1.93 | **2.37** | **1.17-4.80** |
| Maternal smoking during pregnancy | All | 1.36 | 0.80-2.30 | **2.00** | **1.25-3.20** |
|  | Girls | 1.12 | 0.45-2.78 | **2.67** | **1.30-5.51** |
|  | Boys | 1.47 | 0.76-2.84 | 1.68 | 0.89-3.16 |
| Joint exposure to suboptimal gestational factors | All | 1.90 | 0.77-1.52 | **1.37** | **1.01-1.86** |
|  | Girls | 0.88 | 0.48-1.6 | 1.41 | 0.85-2.34 |
|  | Boys | 1.18 | 0.78-1.78 | 1.37 | 0.95-2.06 |

OR= Odds ratio, 95% CI= 95% confidence interval for odds ratio. Multivariate logistic regression models are adjusted for parental education. Significance set at $p < 0.05$ .

**Table 3:** Adjusted multivariate logistic regression model assessing the association between cardiometabolic risk factors and suboptimal gestational factors in QUALITY cohort and boys and girls at age 8-10 years (Visit1)

| Suboptimal gestational condition | Population | Waist circumference ≥ 90<sup>th</sup> percentile | | High density lipoprotein <1.03 mmol/L | |
| --- | --- | --- | --- | --- | --- |
|  |  | OR | 95% CI | OR | 95% CI |
| Gestational diabetes mellitus | All | 1.19 | 0.73-1.92 | 1.18 | 0.73-1.89 |
|  | Girls | 1.34 | 0.62-2.90 | 0.89 | 0.40-1.96 |
|  | Boys | 1.22 | 0.65-2.30 | 1.53 | 0.84-2.79 |
| Hypertensive disorders during pregnancy | All | 1.26 | 0.72-2.18 | 1.40 | 0.82-2.39 |
|  | Girls | 0.44 | 0.16-1.21 | 1.42 | 0.65-3.11 |
|  | Boys | **2.58** | **1.29-5.17** | 1.41 | 0.67-2.96 |
| Maternal smoking during pregnancy | All | **1.96** | **1.24-3.11** | 1.29 | 0.81-2.07 |
|  | Girls | **2.87** | **1.41-5.83** | 1.26 | 0.61-2.59 |
|  | Boys | 1.60 | 0.85-3.01 | 1.40 | 0.74-2.64 |
| Joint exposure to suboptimal gestational factors | All | **1.42** | **1.06-1.91** | 1.32 | 0.99-1.77 |
|  | Girls | 1.33 | 0.82-2.14 | 1.21 | 0.75-1.93 |
|  | Boys | 1.62 | 1.10-2.37 | **1.51** | **1.03-2.22** |

OR= Odds ratio, 95% CI= 95% confidence interval for odd ratio. Multivariate logistic regression models are adjusted for parental education. Significance set at p <0.05.

**Table 4:** Adjusted multivariate logistic regression model assessing the association between anthropometric measures and suboptimal gestational factors in QUALITY cohort and boys and girls at age 10-12 years (Visit 2)

| Suboptimal gestational condition | Population | Overweight |  | Obesity |  |
| --- | --- | --- | --- | --- | --- |
|  |  | OR | 95% CI | OR | 95% CI |
| Gestational diabetes mellitus | All | 0.98 | 0.43-1.86 | 0.59 | 0.26-1.32 |
|  | Girls | 1.15 | 0.38-3.47 | 1.74 | 0.36-3.15 |
|  | Boys | 0.75 | 0.28-2.03 | 0.39 | 0.09-1.77 |
| Hypertensive disorders of pregnancy | All | 0.95 | 0.36-2.47 | 1.05 | 0.43-2.52 |
|  | Girls | 0.20 | 0.02-1.65 | 0.53 | 0.11-2.46 |
|  | Boys | 3.16 | 0.88-11.31 | 1.71 | 0.55-5.29 |
| Maternal smoking during pregnancy | All | 1.30 | 0.60-2.81 | **2.29** | **1.26-4.16** |
|  | Girls | 1.54 | 0.48-4.90 | **2.87** | **1.17-7.03** |
|  | Boys | 1.18 | 0.42-3.32 | 1.90 | 0.85-4.25 |
| Joint exposure to suboptimal gestational factors | All | 0.92 | 0.58-1.46 | **1.63** | **1.15-2.32** |
|  | Girls | 0.97 | 0.48-1.95 | 1.52 | 0.84-2.75 |
|  | Boys | 0.88 | 0.47-1.64 | **1.73** | **1.11-2.71** |

OR= Odds ratio, 95% CI= 95% confidence interval for odd ratio. Multivariate logistic regression models are adjusted for parental education. Significance set at $p < 0.05$ .

**Table 5:** Adjusted multivariate logistic regression model assessing the association between cardiometabolic risk factors and suboptimal gestational factors in QUALITY cohort and boys and girls at age 10-12 years (Visit2)

| Suboptimal gestational condition | Population | Waist circumference ≥ 90<sup>th</sup> percentile |  | High density lipoprotein <1.03 mmol/L |  |
| --- | --- | --- | --- | --- | --- |
|  |  | OR | 95% CI | OR | 95% CI |
| Gestational diabetes mellitus | All | 1.37 | 0.53-3.53 | 0.96 | 0.51-1.80 |
|  | Girls | 1.46 | 0.38-5.55 | 1.24 | 0.49-3.09 |
|  | Boys | 1.42 | 0.36-5.58 | 0.79 | 0.33-1.90 |
| Hypertensive disorders during pregnancy | All | 0.81 | 0.18-3.64 | 0.93 | 0.38-2.29 |
|  | Girls | 0.54 | 0.06-4.6 | 0.45 | 0.09-2.12 |
|  | Boys | 1.30 | 1.15-11.20 | 1.23 | 0.69-2.20 |
| Maternal smoking during pregnancy | All | 2.10 | 0.87-5.08 | 1.71 | 0.95-3.10 |
|  | Girls | 2.78 | 0.86-8.90 | 2.12 | 0.87-5.15 |
|  | Boys | 1.69 | 0.42-6.80 | 1.47 | 0.66-3.26 |
| Joint exposure to suboptimal gestational factors | All | 1.41 | 0.82-2.44 | **1.45** | **1.04-2.03** |
|  | Girls | 1.24 | 0.55-2.76 | 0.71 | 0.42-1.21 |
|  | Boys | 1.73 | 0.81-3.69 | 1.05 | 0.64-1.72 |

OR= Odds ratio, 95% CI= 95% confidence interval for odd ratio. Multivariate logistic regression models are adjusted for parental education. Significance set at p <0.05.

### Stratified analyses : Boys and girls

In boys, at age 8–10 years (V1), hypertensive disorders during pregnancy were positively associated with 137% higher odds of obesity (OR=2.37, 95% CI 1.17-4.80) (Table 2) and 158% higher odds of high waist circumference (OR=2.58, 95% CI 1.29-5.17) (Table 3). Joint exposure to SGFS was positively associated with 51% higher odds of low levels of HDL cholesterol (OR=1.51, 95% CI 1.03-2.22) (Table 3). At age 10–12 years (V2), joint exposure to SGFS was positively associated with 73% higher odds of obesity (OR=1.73, 95% CI 1.11-2.71) (Table 4).

In girls, at age 8–10 years (V1), maternal smoking during pregnancy was positively associated with 167% higher odds of obesity (OR=2.67, 95% CI 1.30-5.51) (Table 2) and 187% higher odds of high waist circumference (OR=2.87, 95% CI 1.41-1.91) (Table 3). At age 10–12 (V2), maternal smoking during pregnancy was positively associated with 187% higher odds of obesity (OR=2.87, 95% CI 1.17-7.03) (Table 4).

## Discussion

A deleterious in-utero environment is suspected of playing a role in cardiometabolic risk factors in childhood^22^ and cardiovascular events in adulthood^23^. The results of the current study reveal that joint exposure to SGFS during pregnancy is positively associated with obesity and an adverse cardiometabolic profile in Caucasian children. Moreover, independent SGFS such as maternal smoking during pregnancy and hypertensive disorders during pregnancy are positively associated with an adverse adipose profile in children. Interestingly, some results appeared to be sex specific and associations persisting with increasing age.

### Joint exposure to SGFS

To the best of our knowledge, the present study is the first to consider joint exposure to SGFS during pregnancy and their associations with childhood overweight, obesity and cardiometabolic risk factors. The pathophysiological mechanisms through which combined SGFS of interest in the present study interact and influence offspring health are still not understood. The current study found that a joint exposure to SGFS was positively associated with a higher odds of obesity, high waist circumference and low levels of HDL cholesterol in the cohort and in boys at age 8–10 years and 10–12 years but not in girls. Taken together, these results suggest that a joint exposure to SGFS results in adverse effects which may be more important in boys than in girls and persist as children age. These findings are consistent with a recent study by Li et al (2017)^24^. This study investigated the impact of GDM on children aged 9–14 years old, and found that boys were more likely to be obese than girls if exposed to GDM during pregnancy. It also suggested that girls display a delayed response to prenatal exposure to GDM, with adverse body weight occurring at the beginning of adulthood. The mechanisms underlying the sex difference in adipose and metabolic outcomes in children are not well understood. Nevertheless, it is possible that boys and girls have different development when exposed to combined SGFS, with girls being more resilient than boys. Murphy and al.’s study on maternal asthma and offspring birth weight also highlighted sex differences. In their study, they showed that boys did not have a reduced birth weight when exposed to mild maternal asthma during pregnancy, while girls had a reduced birth weight^25^. Boys showed signs of compromise only when exposed to an acute maternal asthma exacerbation in utero^26^. These results possibly suggest that boys, contrary to girls, are more sensitive to an adverse gestational environment and this, when the SGFS is exacerbated or is combined with other SGFS, and begin displaying adverse adipose and metabolic profiles during childhood.

### Maternal smoking during pregnancy

Cigarette’s smoking during pregnancy is suspected to cause alterations in the placental barriers structures^27^ leading to later cardiovascular health issues in offspring^28^. In the present study, maternal smoking during pregnancy was associated with an increasing risk of obesity in the cohort and in girls at age 8–10 years and 10–12 years. It was also associated with increasing risk of high waist circumference at age 8–10 years in the cohort. These results therefore suggest that girls are possibly more affected than boys by maternal smoking during pregnancy. Li *et al*. (2016) also found in a Portuguese cohort aged 3–10 years that exposure to maternal smoking during pregnancy increased BMI with age, but no sex specification was given^29^. Shi *et al*. (2013) assessed the potential association of maternal smoking during pregnancy with obesity among Canadian children aged 6–11^30^. They found that it was boys and not girls who were more likely to be obese but did not discuss this sex difference. A possible explanation to our results could be that girls’ early adipose tissue development was more altered than boys’, therefore leading to later obesity. This hypothesis is supported by studies on rats where nicotine altered lipolysis and lipogenesis by inducing a leptin resistance in adipose tissue and liver of female rats, leading to later overweight and metabolic consequences^31^. Another hypothesis justifying the sex difference in this finding could be that the sensitivity in girls and boys toward the cigarette’s components is different. Further studies are yet to be done to investigate these hypotheses considering that in Canada, 20% to 30% of female smokers continue through pregnancy^32^.

### Hypertensive disorders during pregnancy

Hypertensive disorders during pregnancy represent an important worldwide cause of maternal and neonatal mortality ^33^. In high-income countries, gestational hypertension and pre-eclampsia are respectively associated with a 30% and a 3% increased risk of newborn stillbirth^34^. These pregnancy complications are also known to cause growth restriction^35^, a risk factor for later cardiovascular diseases, blood pressure disorders^36^, and a higher BMI in children^37^. In the current study, maternal hypertensive disorders during pregnancy were positively associated with a high waist circumference in boys at age 8–10 (V1). Similar results were found in a systematic review by Davis et al (2012) which included 45,249 children and adolescents and reported that a prenatal exposure to preeclampsia was positively associated with higher odds of higher BMI in both children and adolescents^38^. According to the current literature, the association between hypertensive disorders during pregnancy and childhood obesity goes through a small birth weight for gestational age^39^. Nevertheless, our findings support that the effects of hypertensive disorders during pregnancy potentially go beyond small birth weight for gestational age, given that our statistical analysis found that birthweight was not associated with obesity and cardiometabolic risk factors such as high waist circumference in children. It can be speculated that fetus’s adaptive response to a preeclamptic intrauterine environment results in epigenetic changes that appear later in life, thereby leading to overweight^40^. Another possibility is that heritable genetic factors that predispose to pregnancy hypertensive disorders such as maternal obesity^41^ are inherited by the offspring^22^.

### Strengths and Limitations

This is the first study to examine cross-sectionally the association between adipose profile and cardiometabolic risk factors and three SGFS individually and in combination. It is also reinforced by objective assessment of children’s health. However, the current study was limited by a lack of data on maternal pre-pregnancy BMI and gestational weight gain, both known to have an impact on obesity risk in children^42^ and adolescents^43^. Another limiting factor is that metabolic syndrome criteria are defined starting 10 years of age but we applied them starting at 8 years. The present study also lacked statistical power to analyse cardiometabolic risk factors such as fasting hyperglycemia, and triglycerides due to small sample size.

## Conclusion

The results of this study suggest that intrauterine exposure to maternal smoking during pregnancy, hypertensive disorders during pregnancy and a joint exposure to SGFS (GDM, hypertensive disorders during pregnancy, and maternal smoking during pregnancy) are associated with higher odds of a deleterious adipose and cardiometabolic profiles in Caucasian children. A more marked association of the joint exposure to SGFS was found for boys’ obesity, waist circumference and lipid profile. Moreover, exposure to an independent factor (i.e. maternal smoking during pregnancy, hypertensive disorders during pregnancy) was associated with an adverse adipose profile in children with sex specificities. Consequently, these results suggest that there may be a sex difference in the fetal adaptive mechanisms to a deleterious gestational environment. More research is warranted to explore the key role of intrauterine life on children’s health.

## Data Availability

Data are not available.

## Acknowledgements

Dr. Marie Lambert (July 1952—February 2012), pediatric geneticist and researcher, initiated the QUALITY cohort. Her leadership and devotion to QUALITY will always be remembered and appreciated. The cohort integrates members of TEAM PRODIGY, an inter-university research team including Université de Montréal, Concordia University, Centre INRS—Institut Armand-Frappier, Université Laval, and McGill University. The research team is grateful to all the children and their families who took part in this study, as well as the technicians, research assistants, and coordinators involved in the QUALITY cohort project. We are also grateful to Miguel Chagnon for his help with statistical analyses.

